# Multimorbidity in tuberculosis and its impact on health and treatment outcomes: A prospective cohort study

**DOI:** 10.64898/2026.10.06.26364927

**Authors:** Katherine Jane Hill, Lucy Alinaitwe, Florence Aber, Patricia Nerima, Paul Mubiri, Noela Clara Owarwo, Rogers Owori, Ronald Izati, Wilber Sabiiti, Natasha Walbaum, Molly Naisanga, Stellah Mpagama, Sarah Elizabeth Ellen Mills, Helen Ruth Stagg, Derek James Sloan, Christine Sekaggya-Wiltshire

## Abstract

Multimorbidity – the co-existence of two or more long-term health conditions (LTCs) - is increasingly common in low- and middle-income countries. Yet, in the African region, its prevalence in people with tuberculosis (TB) and its impact on treatment outcomes remain poorly characterised. We sought to determine the prevalence and patterns of multimorbidity and its impact on health and treatment outcomes in people with TB in Kampala, Uganda, and to evaluate cascades of care for hypertension, diabetes mellitus, HIV and mental health conditions.

This was a prospective cohort study of adults (≥18 years) commencing anti-TB treatment at four clinics in Kampala between June 2024–June 2025. LTCs were identified through self-report, validated screening tools, clinical examination and blood analysis, using definitions informed by the Ho *et al.* Delphi consensus. Multimorbidity was defined as TB plus two or more LTCs. Participants were followed to month six to assess health-related quality of life (HRQoL), physiological status (National Early Warning Score (NEWS)), sputum bacillary clearance (TB-Molecular Bacterial Load Assay (TB-MBLA)) and WHO-defined treatment outcomes. Descriptive statistics were used to examine the prevalence and patterns of multimorbidity and evaluate cascades of care. The effect of multimorbidity on health and treatment outcomes was assessed using regression models.

Among 254 adults with TB, the prevalence of multimorbidity was 39.4% (95% CI 33.3– 45.7%); only 22.4% had TB with no co-existing condition. The most prevalent LTCs were HIV (44.1%), undernutrition (39.8%) and depression (16.1%). Care cascades revealed substantial gaps in screening and diagnosis (HIV, diabetes mellitus and mental health conditions) and treatment (hypertension). At enrolment, multimorbidity was associated with worse NEWS (adjusted mean difference 0.88, 95% CI 0.36 to 1.42) and HRQoL (adjusted mean difference in HRQoL score −10.75, 95% CI −16.77 to −4.73). Multimorbidity was associated with a higher risk of a positive sputum TB-MBLA at month 2 and a higher risk of an unsuccessful TB treatment outcome (adjusted risk difference 9.88%, 95% CI 2.59 to 17.17). All deaths occurred among participants with multimorbidity.

Multimorbidity affected two in five adults with TB in this cohort and was associated with worse health at diagnosis and poorer treatment outcomes. National TB programmes are well placed to identify and manage co-existing LTCs, but care cascades show gaps at different stages for different conditions, underscoring the need for tailored, integrated approaches.

## Introduction

Multimorbidity – the co-existence of two or more long-term conditions (LTCs) in an individual - is a rapidly increasing global health challenge [1]. In low- and middle-income countries, the rise of multimorbidity is occurring amidst an ongoing high burden of communicable diseases such as tuberculosis (TB) [2–4]. Shared biopsychosocial determinants and synergism drive the overlap between TB and several LTCs [5, 6]. However, despite calls from the World Health Organization (WHO) in 2022 to consider multimorbidity in people with TB holistically rather than as a series of individual comorbidities, our understanding of multimorbidity in TB is incomplete [7].

Estimates of the prevalence of multimorbidity in people with TB in the African region range from 20.9% - 70.8% [8–10]. Inconsistency in the definitions used and the number of LTCs included likely contributes substantially to this variability, in addition to true differences in underlying epidemiology. An international Delphi consensus study, reported by Ho *et al.*, recommended 24 and 35 LTCs to always and usually include, respectively, in multimorbidity research [11]. While the African region was underrepresented in this study, it remains the leading consensus for international multimorbidity research.

Many TB survivors have persisting morbidity and/or poor health-related quality of life (HRQoL) despite “successful” TB treatment, by WHO-defined treatment outcomes [12–14]. It has been recognised that measuring HRQoL is a crucial component of both care for people with multimorbidity and in determining TB treatment outcomes [12, 14–16].

Uganda is a low-income, high-TB-burden country in East Africa with an increasing burden of non-communicable disease (NCD) [2–4, 17]. No prior study in Uganda has evaluated the prevalence of multimorbidity, applying *Ho et al.* definitions, nor its effect on response to treatment in people with TB. This study primarily sought to determine the prevalence and patterns of individual LTCs and multimorbidity in people at the time of anti-TB treatment (ATT) commencement in the capital city, Kampala. For diabetes mellitus, HIV, hypertension and mental health conditions, we sought to determine the extent to which these were known and managed prior to TB diagnosis. We then examined the effect of multimorbidity on health and clinical characteristics at the start of ATT and on response to ATT.

## Methods

### Study design

This prospective cohort study recruited participants from 6^th^ June 2024 to 13^th^ June 2025 from four TB clinics in central Kampala; the Infectious Diseases Institute – Makerere University (IDI) and three Kampala Capital City Authority (KCCA) TB clinics. Adults (aged ≥18 years) were eligible if they had started ATT for drug-susceptible or drug-resistant TB at any site within the previous 30 days. IDI is a tertiary HIV treatment centre and manages TB solely in people living with HIV (PLHIV), whereas the KCCA clinics manage TB regardless of HIV status.

Study participants were assessed at baseline and at two and six months after ATT commencement, with follow-up assessments conducted by telephone if the participants were unable to attend in person. Study data were collected and managed using Research Electronic Data Capture (REDCap) tools hosted at IDI [18, 19]. Full details of the study design and the assessments conducted can be found in the published protocol [20].

### Sample size

Peltzer *et al.* reported a prevalence of multimorbidity of 20.9% among people with TB in South Africa [8]. This finding, which was the best available estimate at the time of study commencement, was used to inform a precision-based sample size calculation. A sample size of 254 was required to determine the prevalence of multimorbidity to 5% absolute precision.

### Study variables

#### LTCs and multimorbidity

Clinical history at baseline was determined through self-report, structured screening tools, clinical examination and venous blood analysis. The individual LTCs were defined as follows:

- HIV status: Self-report of living with HIV or, if HIV-negative, completion of a negative HIV test within the previous 30 days; otherwise, status was determined by HIV testing. CD4^+^ cell count analysis was performed in PLHIV.
- Nutritional state: Body mass index (BMI) was calculated from height and weight. Undernutrition was defined as a BMI <18.5 kg/m^2^, or a mid-upper arm circumference (MUAC) <23.0cm if participant was pregnant or BMI was missing [4, 21]. Obesity was defined as a BMI ≥30kg/m^2^ [22].
- Alcohol use disorder (AUD): Alcohol Use Disorders Identification Test (AUDIT) score ≥15 or evidence of end-organ alcohol damage and continuing to drink alcohol [23].
- Depression: Patient Health Questionnaire-9 (PHQ-9) score ≥10, or self-reported depression diagnosis [24].
- Anxiety: Generalised Anxiety Disorder-7 (GAD-7) ≥10, or self-reported anxiety diagnosis [25].
- Diabetes mellitus: History of diabetes identified by WHO STEPwise approach to NCD risk factor surveillance tool (STEPS), receiving an anti-diabetic medication, or HbA1c ≥6.5% [26].
- Hypertension: Elevated blood pressure (systolic ≥140mmHg or diastolic ≥90mmHg) at enrolment and month two, one elevated blood pressure reading and history of hypertension identified by WHO STEPS, or receiving an anti-hypertensive medication [27].
- Hepatitis B: Positive hepatitis B surface antigen test [28].
- Severe anaemia: Haemoglobin <8.0 g/dL for men and non-pregnant women, <7.0 g/dL for pregnant women [29].
- Chronic kidney disease: Estimated glomerular filtration rate (eGFR) was calculated from serum creatinine. Chronic kidney disease (CKD) was defined as an eGFR <60 ml/min/1.73m^2^ at enrolment and either on at least one other occasion ≥ two months apart or self-reported CKD [30].
- Peptic ulcer disease (PUD): Self-report of PUD and receiving an antacid medication.
- Cardiovascular disease and cerebrovascular disease: History of either of these conditions identified by WHO STEPS.
- History of other LTCs relied solely on patient self-report of clinical history and current prescriptions.

Multimorbidity, the main analytical variable, was defined as the presence of two or more LTCs in addition to the diagnosis of TB, in accordance with WHO guidance [7]. The Delphi study by Ho *et al*. guided which LTCs were included [11]. Undernutrition, identified by the WHO as a key TB comorbidity, is consistently associated with both increased risk of acquiring TB disease and having a poor outcome [31, 32]. In view of this, and following guidance provided by Ho *et al.*, undernutrition was included in our estimates of multimorbidity. Anaemia – recommended to be usually included in multimorbidity research - is frequently driven by TB disease and uncontrolled HIV. To ensure our multimorbidity estimates focused on multiplicity of healthcare needs for patients and TB clinics, we only included severe anaemia (that would require clinical intervention).

We created two additional multimorbidity variables (‘subtypes of multimorbidity’). Complex multimorbidity was defined as three or more LTCs [33]. Physical-mental multimorbidity was defined as at least one physical LTC and at least one mental health condition or AUD.

Finally, to account for potential reverse causality for both undernutrition and anaemia, we conducted sensitivity analyses with these conditions excluded from the definition of multimorbidity.

#### Measures of health at enrolment

TB-specific HRQoL was assessed using the Functional Assessment of Chronic Illness Therapy – TB (FACIT-TB) questionnaire [34]. As no validated version exists in Luganda - the language most widely spoken in central Uganda - the English version was translated into Luganda by a professional translator for this study. This translation has not been formally validated. The score ranges from 0-180, with higher scores indicating better HRQoL. Physiological health status was assessed by National Early Warning Score (NEWS), calculated from heart rate, respiratory rate, oxygen saturation, requirement for supplementary oxygen, blood pressure, temperature and consciousness level [35, 36].

NEWS ranges from 0-20, with higher scores indicating greater physiological derangement. Participants with pulmonary TB additionally had the following assessments:

- Enrolment sputum bacillary load: Results of sputum Xpert MTB/RIF semi-quantitative bacillary loads were collected from referring clinics and grouped into very low/trace and low/medium/high, which is thought to reflect smear positivity [37].
- Extent of lung involvement: Baseline chest X-rays were reported by two independent clinicians. The presence of cavitation was used in our analyses.

#### Measures of response to ATT

NEWS and HRQoL were measured again at months two and six. Data on the occurrence and severity of adverse drug reactions (ADRs), graded using Common Terminology Criteria for Adverse Events (CTCAE v6.0), were collected [38]. WHO-defined end of ATT outcomes were collected at month six or treatment completion, with TB registers used to identify outcomes for participants lost to study follow-up. Outcomes were categorised as “successful”, “unsuccessful” or “not evaluated”, according to WHO 2021 guidelines [39].

For participants with pulmonary TB, change in sputum bacillary load was assessed using sputum TB-Molecular Bacterial Load Assay (TB-MBLA) analysis. TB-MBLA is a 16S ribosomal RNA-based assay that detects and quantifies viable *Mycobacterium tuberculosis* bacilli in sputum. It has been acknowledged by the WHO as an assay of interest for ATT monitoring and has been shown to be more accurate at characterising response to ATT than Xpert or smear microscopy in Uganda [40, 41]. Sputum TB-MBLA was performed at enrolment and month two and results were used to quantify change in bacillary load and risk of detectable viable mycobacteria at month two.

#### Other variables

Sociodemographic and behavioural data – inclusive of age, sex and smoking status – and details of TB diagnosis were collected at baseline. Socioeconomic status (SES) was determined by the number of markers of poor SES across the following factors: either no household member aged ≥14 years completed at least six years of school or any household member aged 6-18 years not attending school, unemployed (and no employed financial provider) and household crowding (defined as >3 persons per room) [42, 43].

### Statistical analysis

Data were cleaned and analyses performed in R (v4.5.2).

The prevalence and associated 95% confidence intervals (CI) of individual LTCs, multimorbidity, subtypes of multimorbidity and multimorbidity excluding undernutrition and anaemia were estimated using the Clopper-Pearson method and stratified by age, sex and SES.

The effect of multimorbidity on measures of enrolment health and clinical characteristics was assessed by logistic regression for binary outcomes (presence of a cavity on chest X-ray and sputum bacillary load) and Poisson regression for count outcomes (HRQoL and NEWS).

The effect of multimorbidity on response to ATT was assessed by logistic regression for binary outcomes (positive TB-MBLA at month two, any ADR and unsuccessful treatment outcome), proportional odds regression for grade of ADR and Poisson regression for count outcomes (NEWS and HRQoL at months two and six). Two additional analyses were performed for unsuccessful treatment outcome - one using the number of LTCs as the exposure, with a likelihood ratio test (LRT) used as a test of trend, and a sensitivity analysis using that version of the multimorbidity variable.

Negative binomial regression models were used where count outcomes were over-dispersed. All models were adjusted for age, sex, SES and smoking status – the minimal sufficient adjustment set from *a priori* constructed directed acyclic graphs. Marginal standardisation (g-computation) was used to estimate crude and adjusted means and differences.

### Patient and public involvement (PPI)

A core PPI group of three and a PPI panel of eight Ugandan TB survivors were consulted throughout the study. They helped develop the study objectives and case record forms, aided recruitment and guided interpretation and dissemination of study results.

### Ethical approval

Ethical approval was prospectively obtained from IDI Research Ethics Committee (on 23/02/2024, IDI-REC-2023-82), the University of St Andrews School of Medicine Ethics Committee (on 04/04/2024, MD17720) and the Uganda National Council of Science and Technology (on 07/05/2024, HS3888ES). Written informed consent was obtained from all participants, and the study was conducted according to good clinical practice principles.

## Results

Two hundred and fifty-four participants with active TB were enrolled; 79/254 (31%) were female and the baseline median age was 35 years (interquartile range (IQR) 27-42 years) (Table 1). Most participants had pulmonary TB (241/254, 95%). Only one participant (0.4%) had drug-resistant TB.

**Table 1:**
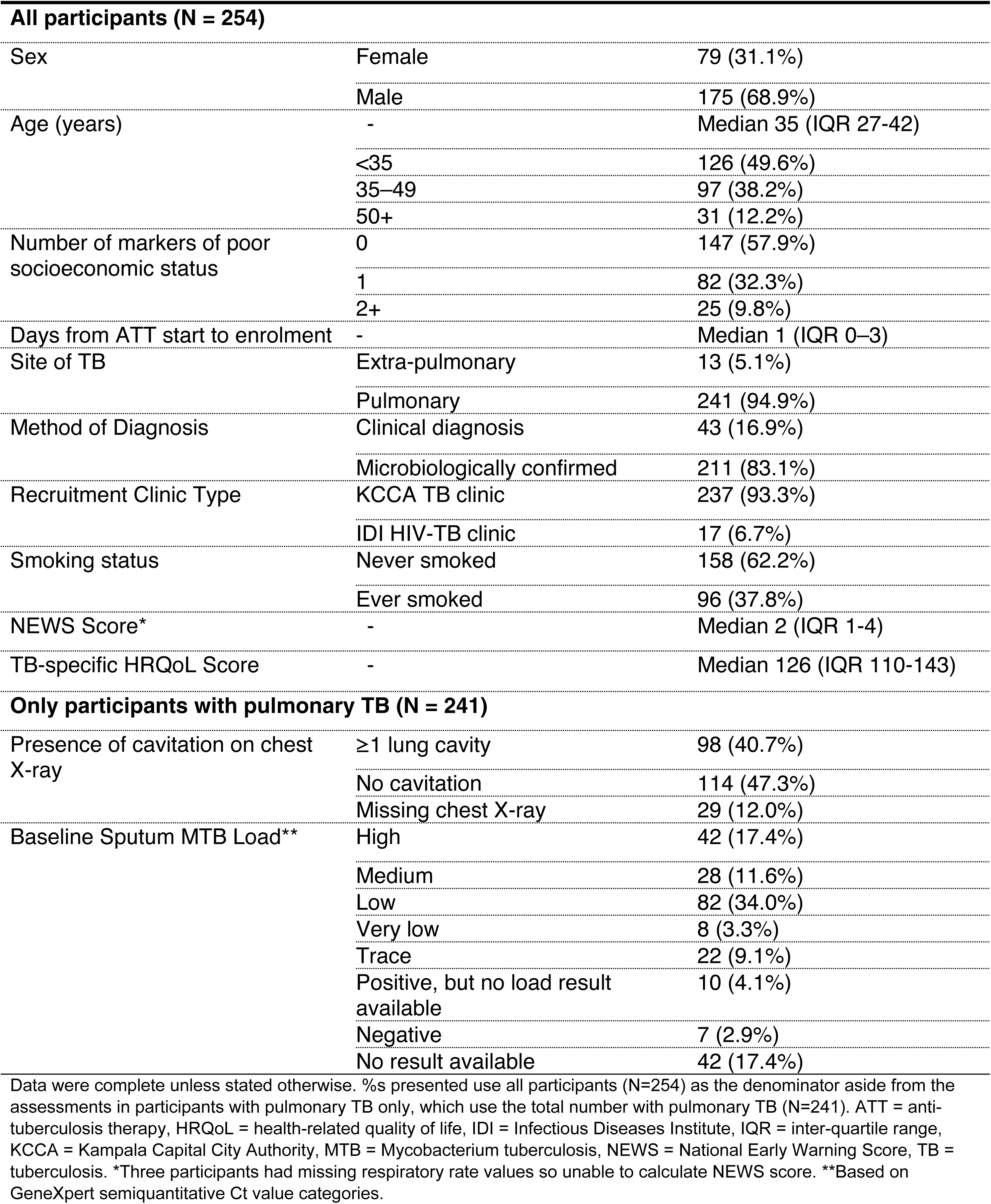
Characteristics of the 254 participants included in the cohort at enrolment.

### Prevalence and patterns of multimorbidity

The overall prevalence of multimorbidity was 39.4% (95% confidence interval (CI): 33.3-45.7%, 100/254) at enrolment. In the sensitivity analysis, with anaemia and undernutrition excluded, 23.6% (95% CI: 18.5-29.3%, 60/254) had multimorbidity (Fig A in S1 Appendix). The prevalence estimates of multimorbidity were higher in females than males and increased with increasing age and number of markers of poor socioeconomic status, although confidence intervals were overlapping (Fig 1). Using the multimorbidity subtype definitions, 13.4% (95% CI: 9.5-18.2%, 34/254) had complex multimorbidity and 17.7% (95% CI: 13.2-23.0%, 45/254) had physical-mental multimorbidity.

**Fig 1.**
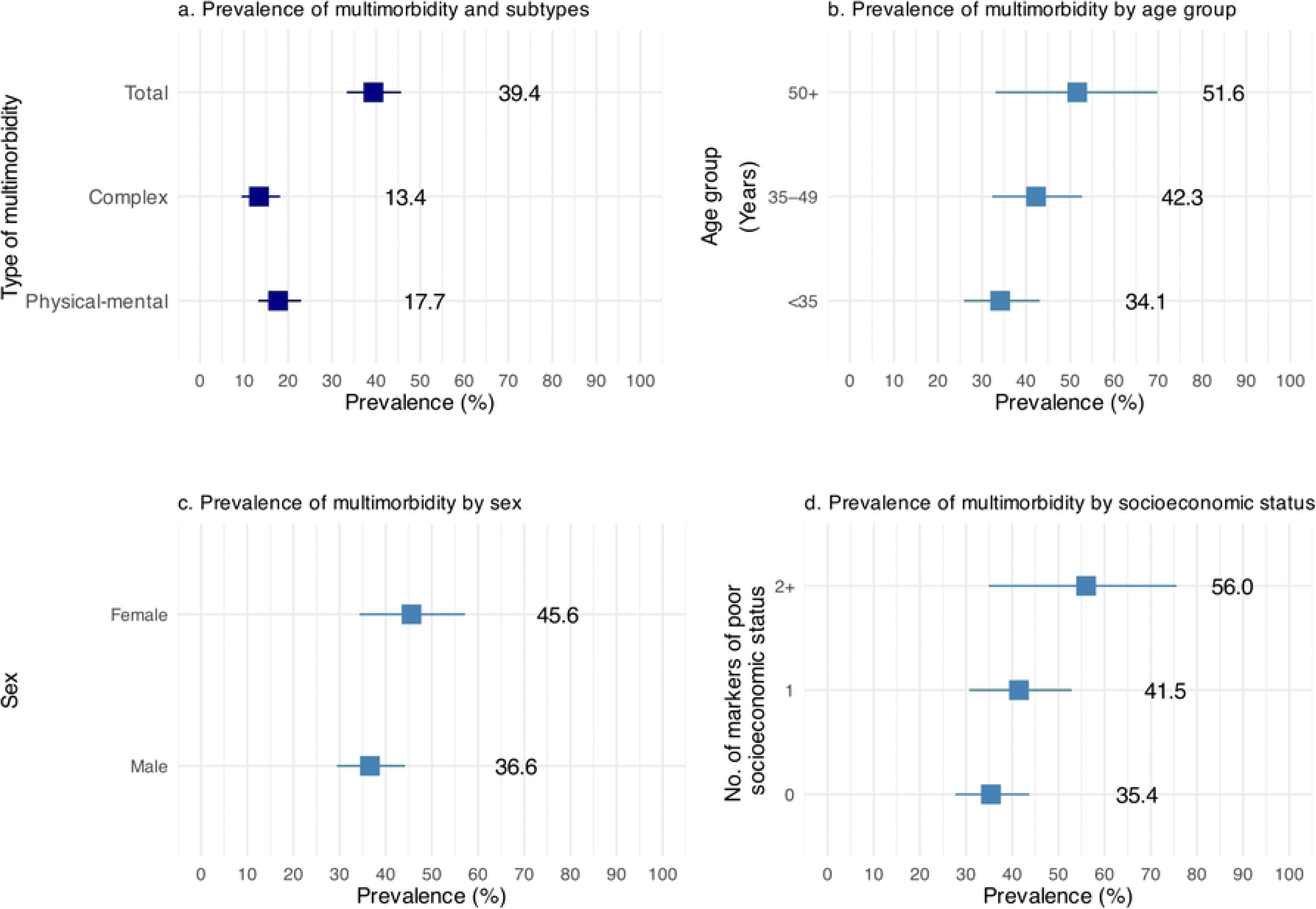
Prevalence of multimorbidity. Prevalence (boxes, numbers) and 95% confidence intervals (lines) of multimorbidity at enrolment among the cohort of participants. Panel a demonstrates the prevalence of multimorbidity (‘total’) and subtypes of multimorbidity (complex and physical-mental). Panels b-d demonstrate the prevalence of multimorbidity by age group, sex and socioeconomic status, respectively, using the main variable.

The most prevalent individual health conditions were HIV (112/254, 44.1%), undernutrition (101/254, 39.8%), depression (41/254, 16.1%), diabetes mellitus (19/254, 7.5%) and alcohol use disorder (17/254, 6.7%) (Table B and Fig C in S1 Appendix). Only 22.4% (57/254) participants had TB and no co-existing conditions (Fig 2).

**Fig 2.**
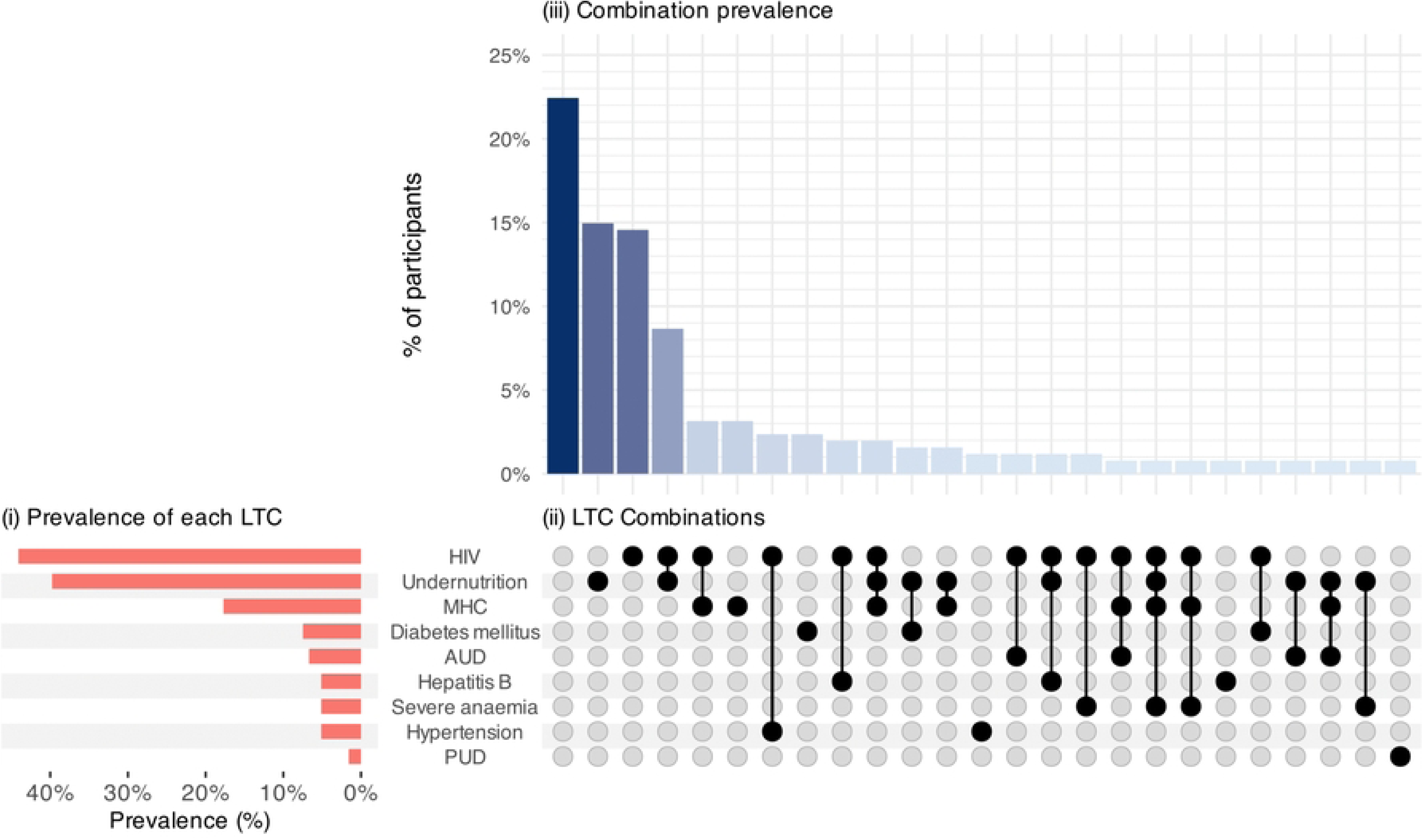
Prevalence and intersections of long-term conditions. Three panels documenting the prevalence of individual and combinations of long-term conditions (LTCs) in the cohort. (i) The set size plot (bottom left panel) shows the prevalence of each LTC. (ii) The matrix plot (bottom right panel) shows the combinations of LTCs; filled circles indicate the LTCs present in each combination, ordered by frequency. (iii) The intersection plot (top right panel) shows the percentage of participants with each LTC combination represented in the matrix plot. For example, at enrolment 9% of people were living with HIV and had undernutrition but no other LTCs. Percentages are calculated using the full cohort as the denominator (N=254). Intersections are mutually exclusive; each participant contributes to one combination only. AUD = alcohol use disorder, HIV = human immunodeficiency virus, LTC = long-term condition, MHC = mental health condition (depression, anxiety or other mental health condition, but excluding alcohol use disorder), PUD = peptic ulcer disease.

**Fig 3.**
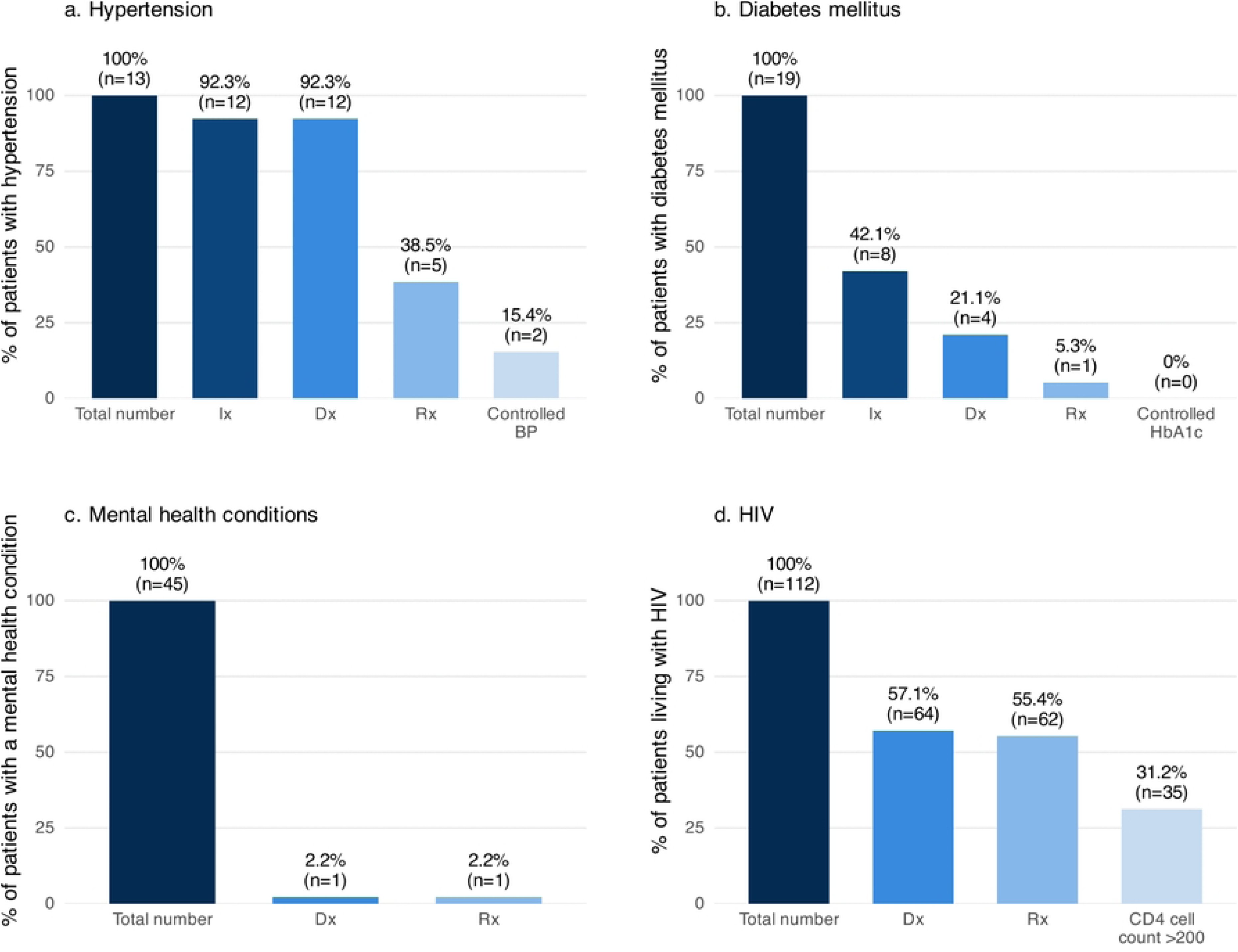
Cascades of care for participants living with hypertension, diabetes mellitus, mental health conditions and HIV. Bar charts representing numbers and percentages of participants (out of the total number of participants with each long-term condition (LTC)) who were previously investigated (hypertension and diabetes mellitus only), previously diagnosed and receiving treatment for a. hypertension, b. diabetes mellitus, c. mental health conditions (depression, anxiety or other mental health conditions, but excluding alcohol use disorder) and d. HIV. Percentage with controlled disease also presented for hypertension (defined as a systolic blood pressure <140mmHg and diastolic blood pressure <90mmHg) and diabetes mellitus (defined as a glycated haemoglobin (HbA1c) <7.0%). Proportion of participants with a CD4^+^ cell count > 200 cells/µL presented for people living with HIV. BP = blood pressure, Dx = previously diagnosed, Ix = previously screened, Rx = receiving treatment for the LTC.

### Cascades of Care

Among participants with hypertension, 12/13 (92.3%) had previously had their blood pressure measured and received a diagnosis of hypertension (**Error! Not a valid bookmark self-reference.**). However, only 5/13 (38.5%) were receiving anti-hypertensive treatment, and only 2/13 (15.4%) had controlled blood pressure. In contrast, for diabetes mellitus and mental health conditions, attrition occurred earlier in the care cascade. Fewer than half of participants with diabetes had previously had their blood glucose measured (8/19, 42.1%), and only 4/19 (21.1%) had received a prior diagnosis of diabetes. Only 1/45 (2.2%) of participants with a mental health condition had a prior diagnosis. Among the 112 PLHIV, only 64 (57.1%) were diagnosed with HIV prior to presenting with TB. However, almost all those known to be living with HIV prior to TB were receiving anti-retroviral therapy (62/64, 96.9%).

### Effect of multimorbidity on clinical characteristics at enrolment

Participants with multimorbidity had worse markers of physiological health and HRQoL at enrolment. NEWS scores for participants with multimorbidity were higher by 0.78 points (95% CI 0.27-1.29) than for those without multimorbidity (Table D in S1 Appendix). After adjustment, this difference increased to 0.88 points (95% CI 0.36-1.42, p-value <0.01). Mean total HRQoL scores were 10.02 points lower (95% CI -15.97 to -4.08), 10.75 points after adjustment (95% CI -16.77 to -4.73, p-value <0.01) (S1 Appendix). When studying different domains of HRQoL, no evidence of an association was found between multimorbidity and spiritual and social domains, but participants with multimorbidity had reduced physical (adjusted mean difference (aMD) -5.42, 95% CI -8.66 to -2.18, p <0.01), emotional (aMD -3.78, 95% CI -5.89 to -1.67, p<0.01) and functional domains (aMD -1.57, 95% CI -3.04 to - 0.11, p=0.04) (Table E in S1 Appendix).

No evidence of an association was demonstrated between multimorbidity and presence of lung cavitation or sputum bacillary burden at enrolment (S1 Appendix).

### Effect of multimorbidity on response to ATT

At month two and month six, respectively, 215 and 210 participants remained under study follow-up (Fig F and Table G in S1 Appendix).

Programmatic TB treatment outcomes were obtained for all participants, although two were recorded as “not evaluated” and excluded from the analysis. A WHO-defined successful treatment outcome was recorded in 236/254 (92.9%) of participants. Among the 16 (6.3%) with an unsuccessful outcome, 6/254 (2.4%) died, 2/254 (0.8%) failed treatment and 8/254 (3.1%) were lost to follow-up. Participants with multimorbidity had an increased risk of an unsuccessful treatment outcome; the crude risk difference (RD) was 9.51% (95% CI 2.60-16.42) and aRD was 9.88% (95% CI 2.59-17.17, p-value <0.01) (Table 2). Notably, all participants who died (causes described in Table H in S1 Appendix) had multimorbidity at baseline. Risk of an unsuccessful outcome increased as the number of LTCs increased (LRT for trend p-value <0.01) (Table 2). On sensitivity analysis, a direction of effect towards increased risk of an unsuccessful treatment outcome with multimorbidity remained even when undernutrition and anaemia were excluded from the multimorbidity definition, but confidence intervals were wider and included the null (aRD 6.95%, 95% CI -1.90-15.80, p=0.08) (Table J in S1 Appendix).

**Table 2:** Results of regression analyses on the effect of multimorbidity on risk of an unsuccessful TB treatment outcome.

| Variable | Number<br>(n(%)) | Crude |  |  | Adjusted |  |  |
| --- | --- | --- | --- | --- | --- | --- | --- |
|  |  | Marginal Risk (%) | RD (95% CI) (%) | p-value | Marginal Risk (%) | RD (95% CI) (%) | p-value |
| Multimorbidity |  |  |  |  |  |  |  |
| No MM | 153 (60.7) | 2.61 | Ref | p=0.003 | 2.57 | Ref | p=0.003 |
| MM | 99 (39.3) | 12.12 | 9.51<br>(2.60-16.42) |  | 12.45 | 9.88<br>(2.59-17.17) |  |

**b. The effect of number of LTCs on risk of an unsuccessful TB treatment outcome**
| Variable | Number<br>(n(%)) | Crude |  |  | Adjusted |  |  |
| --- | --- | --- | --- | --- | --- | --- | --- |
|  |  | Marginal Risk<br>(%) | RD (95% CI) (%) | p-value | Marginal Risk<br>(%) | RD (95% CI) (%) | p-value |
| Number of LTCs |  |  |  |  |  |  |  |
| 0 | 56<br>(22.2) | 1.79 | Ref | p for<br>trend<br>=0.003 | 1.79 | Ref | p for<br>trend<br>=0.003 |
| 1 | 97<br>(38.5) | 3.09 | 1.31 (-3.58-6.20) |  | 3.00 | 1.21 (-3.62-6.04) |  |
| 2+ | 99<br>(39.3) | 12.12 | 10.34 (3.03-17.64) |  | 12.48 | 10.69 (2.96-18.43) |  |

Tables showing the effect of multimorbidity (a) or number of co-existing long-term conditions (b) on risk of an unsuccessful TB treatment outcome, as defined by the WHO. Participants with an outcome of "not evaluated" were excluded. Models are adjusted for age, sex, history of smoking and number of markers of poor socioeconomic status. p-values are from likelihood ratio tests (LRT). In panel b, the p-values are LRTs for linear trend. CI = confidence interval, LTC = long-term condition, MM = multimorbidity, RD = risk difference, Ref = reference, TB = tuberculosis.

Sputum TB-MBLA analysis was performed in 103 participants at month two, 18 of whom (17.5%) had detectable *M. tuberculosis*. Multimorbidity was associated with a higher risk of a positive month two TB-MBLA; aRD 19.1% (95% CI 1.1-37.0%, p-value 0.03) (Table K in S1 Appendix). Only 57 participants had both a positive sputum TB-MBLA at enrolment and repeat TB-MBLA analysis at month two. We were therefore unable to complete our planned analysis on the effect of multimorbidity on change in sputum bacillary burden.

NEWS and HRQoL improved through treatment for both those with and without multimorbidity (Fig 4). By month six, there was no evidence of a difference in HRQoL by multimorbidity, but mean NEWS remained worse in participants with multimorbidity (aMD 0.45, 95% CI 0.11-0.79, p-value <0.01) (Table L in S1 Appendix).

**Fig 4.**
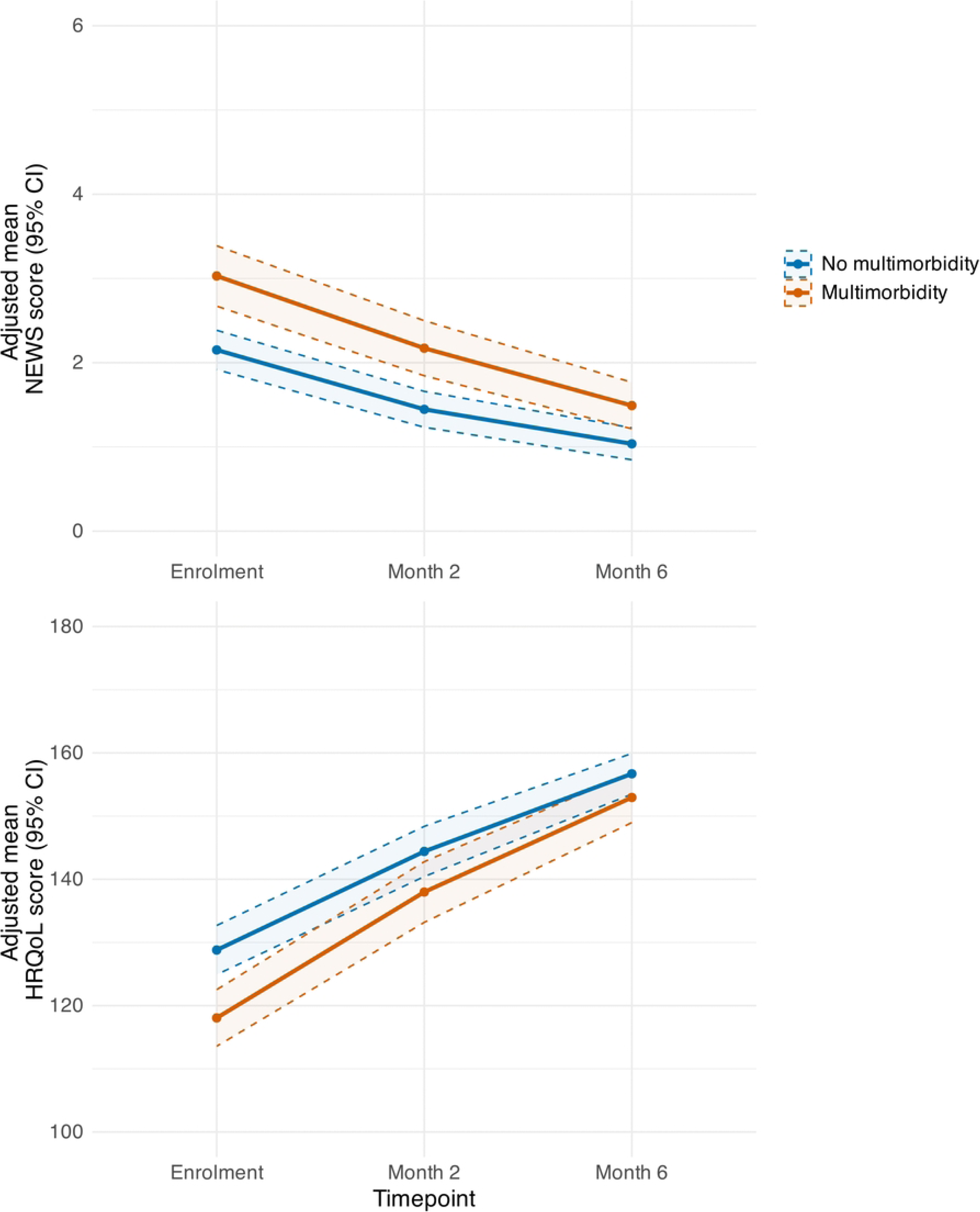
Change in early warning scores and health-related quality of life through TB treatment. Solid line represents mean scores, dotted lines and shadows represent 95% confidence intervals. Means and confidence intervals are calculated by marginal standardisation from regression model for the effect of multimorbidity on outcome (either NEWS score or HRQoL score), adjusted for age, sex, socioeconomic status and smoking status. Red colour represents participants with multimorbidity, blue colour represents participants without multimorbidity. Possible scores range from 0 to 20 for NEWS and 0 to 180 for HRQoL score, but y-axes truncated for clarity. CI = confidence interval, NEWS = National Early Warning Score, HRQoL = TB-specific health-related quality of life score.

Seventy-four participants (74/210, 35.2%) had at least one ADR during treatment (Table H in S1 Appendix). There was a direction of effect towards higher risk of having an ADR (aRD 9.23%, 95% CI -3.87-22.32, p=0.16) and higher odds of having a more severe ADR in participants with multimorbidity (adjusted odds ratio 1.66, 95% CI 0.90-3.06, p=0.10), but both estimates had wide confidence intervals inclusive of the null (Table M in S1 Appendix).

## Discussion

Multimorbidity is common in adults commencing treatment for TB disease; 39.4% of this cohort had two or more LTCs, whilst only 22.4% had TB without any co-existing health conditions. Heterogeneity in definitions of LTCs and multimorbidity in TB has contributed to inconsistent and heterogeneous estimates of TB-multimorbidity across Africa. The WHO defines TB-multimorbidity as “*the presence of two or more concurrent diseases or health conditions in a person with TB*” [7]. Prior studies which reported higher prevalences of TB-multimorbidity defined it as TB plus one or more concurrent diseases [9]. Variability in numbers of LTCs measured and inclusion of behavioural risk factors into multimorbidity estimates have further contributed to variation in reported prevalences [8, 10, 44]. By conducting prospectively-collected, detailed LTC assessments we have been able to apply internationally agreed definitions of LTCs and multimorbidity more closely and provide a detailed assessment of the burden and patterns of TB-multimorbidity [7, 11]. Nevertheless, differences in definitions should not obscure the overall message emerging from research to date: people with TB in Africa present with multiple complex healthcare needs.

Multimorbidity in this cohort was associated with worse health at TB diagnosis, reflected in both higher NEWS and worse HRQoL. We did not show an effect on sputum bacillary burden or chest X-ray findings. HIV, which 75.0% of participants with multimorbidity were living with, is associated with paucibacillary disease and fewer chest X-ray changes [45, 46]. Yet a reverse relationship is seen for other LTCs, such as undernutrition and diabetes where increased bacillary burden and more extensive radiological changes are seen [47–50]. Our cohort was too small to allow disaggregation of bacillary burden or radiological changes by HIV status. These conventional markers of TB severity at diagnosis may be less reliable in patients with multimorbidity and this emphasises the importance of holistic clinical assessment and use of patient-reported measures of health. Additionally, it is important that we consider individual disease mechanisms in multimorbidity research and clinical care.

Participants with multimorbidity had an increased risk of an unsuccessful TB treatment outcome, the risk increased as the number of co-existing LTCs increased and all deaths were in participants with multimorbidity. This association was demonstrated despite the treatment success rate in this study (93%) being higher than programmatic treatment success in Kampala (89%) [51], and participants with multimorbidity benefitting from screening and referral for management of LTCs as part of this study. The deleterious effect of multimorbidity on response to ATT was further supported by the evidence of an increased risk of viable TB remaining in sputum at two months on TB-MBLA testing. WHO TB treatment outcomes and bacteriological analysis are crude measures of response to ATT and do not capture sequelae or long-term consequences post-TB. Future studies examining the consequences of multimorbidity in people with TB should also look at health and functional status beyond the end of treatment, including post-TB sequelae, TB recurrence and early mortality from other causes.

These findings starkly highlight the need for national TB programmes (NTPs) to better integrate care for comorbidities into TB services, an approach currently under review by the Ugandan NTP. However, the cascades of care analyses suggest that gaps in care are falling at different stages for different LTCs. Only 57.1% of PLHIV in this cohort knew their status prior to presenting with TB disease, which falls well below the UNAIDS target of 95% for all PLHIV [52]. Continued work to identify PLHIV prior to presentation with TB, which is a marker of advanced HIV disease, is required. However, once diagnosed, linkage to care and commencement of anti-retroviral treatment was good in this cohort (62/64 PLHIV, 96.9%).

Most participants with hypertension knew their diagnosis prior to TB and instead, improvement in treatment and control is required, as also shown in Ugandan studies in the general population [53, 54]. For diabetes and mental health conditions, the gaps in care are at the point of screening (for diabetes) and diagnosis. Engagement in TB clinics, especially in settings with underdeveloped primary care and high patient costs associated with attending health clinics, provides a unique opportunity to identify and facilitate management of additional healthcare needs [55]. TB and HIV programmes have spearheaded integrated care, and the co-management of these conditions is estimated to have saved nine million lives between 2005-2020 [7]. Consideration should be given to how this approach could be adapted and applied to a wider range of LTCs. However, the optimal timing of such interventions also needs to be carefully considered. Screening for LTCs and linking to care at the point of TB diagnosis, as seen in a multi-site study across East and Southern Africa, reduces persistence of symptoms and disability at the end of ATT [56]. However, diagnostic tools (such as blood pressures, serum markers of disease and mental health questionnaires) can be directly impacted by acute illness, risking under- or over-diagnosis, and patients may already be overwhelmed at TB diagnosis without additional diagnoses and medications. Yet delaying interventions misses the opportunity to potentially impact treatment outcomes and fails patients who are lost to follow-up.

### Strengths and limitations

There are several strengths of this study. Screening for a broad range of LTCs, directed by Ho *et al.,* and using validated screening tools provides a detailed analysis of multimorbidity at TB diagnosis. The use of objective and patient-reported measures of clinical characteristics and health at TB diagnosis and throughout TB treatment has allowed a thorough description of the various facets of effects of multimorbidity. PPI from study conception onwards has ensured that the study is relevant to people with TB in Uganda. There remain some limitations, however. Whilst the emphasis on cross-sectional diagnoses of LTCs at enrolment mitigated survivor bias, our estimates of some LTCs may be imprecise. The prevalence of hypertension may be underestimated due to acute illness hypotension at the start of TB treatment. We used HbA1c instead of serum blood glucose measurements in our diabetes screening to mitigate overdiagnosis due to transient TB-induced hyperglycaemia. Yet, despite this, diabetes prevalence was likely still overestimated and glycaemic control underestimated. Similarly, defining depression and anxiety using screening tools administered at a single time point may overestimate their prevalence, particularly for depression, whose symptoms can overlap with those of acute TB [57].

Although FACIT-TB was professionally translated into Luganda, the lack of validation may limit comparability with other studies using FACIT-TB. The percentage of patients with extra-pulmonary TB in this cohort was lower than prior estimates of 12.9%, which may limit generalisability of our findings [58]. Recruitment solely from outpatient clinics may have missed the most unwell patients, leading to underestimation of the prevalence of multimorbidity in the total TB population.

### Conclusions

In this cohort of adults with TB in Uganda, we found a high prevalence of multimorbidity at the start of TB treatment. Multimorbidity was associated with worse health and HRQoL at TB diagnosis and a higher risk of an unsuccessful TB treatment outcome. Screening for and management of LTCs should be integrated into NTPs, which are uniquely positioned to care for people with multimorbidity.

## Data Availability

The data underlying this study contain sensitive health information, including HIV status and mental health conditions, collected from a small cohort at identifiable clinic sites. Participants could potentially be re-identified, so the data cannot be shared publicly. Under the study's data management agreement, pseudo-anonymised data are held by the Infectious Diseases Institute (IDI), Makerere University, as data controller. Researchers who meet the criteria for access to confidential data can request it through IDI's established Standard Operating Procedures for data access, by contacting IDI's research department. Analysis code will be made available on publication at GitHub (https://github.com/khill280/MITICare-analysis.git)

## Acknowledgements

We are grateful to the personnel from the Infectious Diseases Institute and KCCA TB clinics who supported this study, the patient and public involvement members who contributed throughout this study to its success, and to all the people who participated in the study.

## Supporting information

**S1 Checklist. STROBE checklist.**

**S1 Appendix. Supplementary material.**

